# Correlation between umbilical cord torsion and Doppler parameters of umbilical artery: a single-center retrospective case-control study

**DOI:** 10.1101/2023.11.12.23298437

**Authors:** Yuan Li, Dirong Zhang, Yu Shi

**Affiliations:** Department of Ultrasound, Peking University Shenzhen Hospital, Shenzhen, Guangdong, 518036, China; Shenzhen Key Laboratory for Drug Addiction and Medication Safety, The Hong Kong University of Science and Technology Medical Center, Shenzhen, Guangdong, 518036, China

**Keywords:** Prenatal ultrasound assessment, Cord torsion, Umbilical artery, Color Doppler ultrasound

## Abstract

**Objective:** To explore the correlation between umbilical cord torsion and the changes of umbilical artery Doppler parameters, to provide valuable information for prenatal ultrasound screening of umbilical cord torsion, and to explore the possible mechanism of umbilical artery Doppler parameters changes during umbilical cord torsion. Method:The subjects were 962 pregnant women who were discharged from our hospital from January 2015 to November 2021 and were eligible for inclusion (415 in the case group and 547 in the control group). The measurement data of umbilical artery Doppler parameters (PSV, S/D, RI, PI) were collected from 21 to 40 weeks of gestation, and the differences among the above parameters were statistically analyzed.

**Results:** The peak systolic velocity (PSV) of umbilical artery is positively correlated with gestational age, while the Doppler resistance parameters (S/D, RI, PI) of umbilical artery are negatively correlated with gestational age. The mean values of umbilical artery Doppler parameters (PSV, S/D, RI, PI) in the case group were significantly lower than those in the control group of the same pregnancy age (P < 0.05).

**Conclusion:** The decrease of Doppler parameters of umbilical artery in late pregnancy is significantly related to umbilical cord torsion, which may be a clue for prenatal ultrasound screening of umbilical cord torsion. The Doppler parameters of umbilical artery during umbilical cord torsion are consistent with the basic principle of Doppler parameters change after vascular stenosis in other parts of the human body. It is necessary to conduct multicenter prospective study in the future.

## 1. Introduction

The umbilical cord is very important for the normal development, survival and health of the fetus after birth. The proper helix of umbilical cord and its tissue structure provide some pressure protection for umbilical vessels^[1-2]^. The physiological torsion of umbilical cord can reach 6-11 weeks, and the torsion of umbilical cord can be more than 12 weeks^[3]^. Torsion of umbilical cord may lead to fetal growth restriction, premature delivery, meconium contamination of amniotic fluid, and even stillbirth^[4-8]^. Umbilical cord torsion as one of the causes of perinatal fetal death^[9]^. Prenatal ultrasound has been exploring the ultrasonic manifestation of umbilical cord torsion, but no consensus has been reached^[10,11]^. At present, there is no study on the relationship between umbilical cord torsion and the changes of umbilical artery Doppler parameters. The purpose of this study is to explore the correlation between umbilical cord torsion and the changes of umbilical artery Doppler parameters, to provide valuable information for prenatal ultrasound screening of umbilical cord torsion, and to explore the possible mechanism of umbilical artery Doppler parameters changes during umbilical cord torsion.

## 2. Materials and Methods

This study is a retrospective pathological control study. Pregnant women who underwent routine prenatal ultrasound examination and hospital delivery in our hospital from January 2015 to November 2021 were randomly collected.

### Inclusion criteria

1. Pregnant women who give birth in our hospital have definite discharge diagnosis or pathological diagnosis of placental appendages;
2. Low-risk pregnancy, including no pregnancy complications or major underlying diseases, such as gestational hypertension, gestational diabetes, placental previa, placental abruptio, etc.;
3. There is a record of prenatal ultrasound examination in our hospital, and the Doppler parameters of umbilical artery are measured in the free segment of umbilical cord during the examination;
4. Singleton pregnancy.

### Exclusion criteria

1. Severe fetal malformations or chromosome abnormalities;
2. Pregnant women with single umbilical cord artery or other abnormal umbilical cord blood vessels;
3. Stillbirth or induced labor due to other reasons.

Gestational age was calculated based on last menstrual period. If the last menstrual period is unclear, it is calculated and recorded according to the NT cycle after correcting the gestational age. Obstetricians diagnosed the umbilical cord after delivery, and defined those who were discharged as having umbilical cord torsion as the case group, and those who were diagnosed as having no umbilical cord torsion were randomly selected as the control group. The ethics committee of our hospital agreed to waive informed consent for this study.

The ultrasonic instrument uses GE and Mindray color Doppler ultrasound diagnostic machine and the probe frequency is 4∼8MHz. The free umbilical cord floating in the amniotic fluid was measured by Doppler, and the measurement Angle was as parallel as possible to the direction of umbilical cord blood flow. Umbilical artery Doppler blood flow parameters, including peak systolic velocity (PSV), umbilical artery blood systolic/diastolic ratio (S/D), resistance index (RI) and pulse index (PI), were extracted from the workstation at each gestational week from 21 to 40 weeks.

SPSS26.0 statistical software was used for data processing:1. All umbilical artery Doppler blood flow parameters in the case group and the control group at 21-40 weeks gestation were counted according to gestational age. The mean of each gestational age measurement data in the two groups was calculated; 2. The change trend of umbilical artery blood flow parameters with gestational week was analyzed.3. Logistic regression analysis was performed on the umbilical artery blood flow parameters of the case group and the control group of the same gestational age, P < 0.05, indicating significant differences between the two variables. Regression analysis fills in the missing values.

## 3. Results

This study included 1608 pregnant women who delivered in our hospital (666 in the case group and 942 in the control group). According to the inclusion exclusion criteria, 646 pregnant women (251 in the case group and 395 in the control group) were excluded, and 415 pregnant women in the case group and 547 pregnant women in the control group were finally included. Pregnant women ranged in age from 21 to 44 years, with a mean age of 31 years (P > 0.05). A total of 4215 umbilical artery Doppler flow parameters were collected in the case group (1676 in the case group and 2543 in the control group), of which basically each included the peak rate of umbilical artery contraction (PSV) and the umbilical artery flow resistance parameters (S/D), and some of them lacked the resistance parameters RI (19) and PI (75) in the umbilical artery group. The average values of umbilical artery Doppler parameters in the case group and the control group at different gestational ages were shown in Table 1, indicating that the average values of PSV, S/D, RI and PI of umbilical artery in the case group were lower than those in the control group at different gestational ages.

**Table 1.** Mean values of umbilical artery PSV, S/D, RI and PI between case groups and control groups at different gestational weeks.

|  |  |  |  |  |  |  |  |  |  |  |  |  |
| --- | --- | --- | --- | --- | --- | --- | --- | --- | --- | --- | --- | --- |
| GA | Number |  | S/D |  | RI |  | PI |  | PSV |  |  |  |
|  | Case | Control | Case | Control | Case | Control | Case | Control | Case | Control | Case | Control |
|  | group | group | group | group | group | group | group | group | group | group | group | group |
| 21 | 33 | 94 | 3.09 | 3.43 | 0.66 | 0.70 | 1.07 | 1.18 | 33.22 | 35.1 |  |  |
| 22 | 102 | 183 | 3.17 | 3.21 | 0.67 | 0.68 | 1.1 | 1.11 | 35.38 | 35.06 |  |  |
| 23 | 82 | 95 | 3.05 | 3.14 | 0.66 | 0.67 | 1.07 | 1.09 | 35.66 | 36.45 |  |  |
| 24 | 25 | 37 | 2.92 | 3.06 | 0.65 | 0.66 | 1.04 | 1.09 | 36.04 | 36.62 |  |  |
| 25 | 9 | 51 | 2.6 | 2.99 | 0.62 | 0.65 | 0.91 | 1.03 | 37.18 | 40.81 |  |  |
| 26 | 10 | 54 | 2.61 | 2.95 | 0.60 | 0.65 | 0.93 | 1.03 | 44.48 | 42.49 |  |  |
| 27 | 17 | 28 | 2.44 | 2.77 | 0.58 | 0.62 | 0.86 | 0.96 | 37.99 | 42.61 |  |  |
| 28 | 35 | 49 | 2.46 | 2.73 | 0.59 | 0.62 | 0.85 | 0.96 | 41.2 | 45.01 |  |  |
| 29 | 47 | 80 | 2.45 | 2.67 | 0.58 | 0.61 | 0.86 | 0.92 | 42.33 | 46.14 |  |  |
| 30 | 101 | 172 | 2.39 | 2.64 | 0.57 | 0.60 | 0.85 | 0.91 | 44.84 | 47.01 |  |  |
| 31 | 111 | 153 | 2.4 | 2.57 | 0.57 | 0.60 | 0.84 | 0.89 | 43.47 | 47.09 |  |  |
| 32 | 97 | 132 | 2.36 | 2.54 | 0.56 | 0.59 | 0.82 | 0.89 | 42.81 | 47.79 |  |  |
| 33 | 50 | 71 | 2.27 | 2.44 | 0.54 | 0.58 | 0.81 | 0.87 | 41.36 | 48.74 |  |  |
| 34 | 47 | 60 | 2.12 | 2.43 | 0.52 | 0.57 | 0.73 | 0.85 | 44.08 | 49.16 |  |  |
| 35 | 54 | 69 | 2.18 | 2.39 | 0.53 | 0.56 | 0.76 | 0.85 | 45.76 | 49.64 |  |  |
| 36 | 128 | 212 | 2.14 | 2.31 | 0.52 | 0.55 | 0.75 | 0.83 | 46.91 | 50.3 |  |  |
| 37 | 243 | 321 | 2.13 | 2.27 | 0.52 | 0.54 | 0.75 | 0.8 | 46.38 | 50.58 |  |  |
| 38 | 183 | 279 | 2.11 | 2.25 | 0.52 | 0.54 | 0.73 | 0.8 | 46.59 | 50.78 |  |  |
| 39 | 202 | 293 | 2.07 | 2.2 | 0.50 | 0.53 | 0.73 | 0.78 | 45.79 | 51 |  |  |
| 40 | 96 | 110 | 2.06 | 2.17 | 0.50 | 0.53 | 0.72 | 0.78 | 47.16 | 51.13 |  |  |
| GA: gestational age; PSV :peak systolic velocity; S/D: systolic/diastolic ratio; RI: resistance index; PI: pulsatility index |  |  |  |  |  |  |  |  |  |  |  |  |

The correlation analysis of umbilical artery flow parameters (PSV, S/D, RI, PI) and gestational age in case group and control group respectively showed that the peak velocity of umbilical artery contraction (PSV) was positively correlated with gestational age in both groups, while the correlation parameters of umbilical artery Doppler resistance (S/D, RI, PI) were negatively correlated with gestational age in both groups(R_PSV_ 0.374 vs 0.538,R_S/D_ 0.617 vs 0.619,R_RI_ 0.616 vs 0.563,R_PI_ 0.602 vs 0.622,P>0.05)○

After filling in the missing values by regression analysis, Logistic regression analysis showed that:In weeks 22-27, the umbilical artery Doppler parameters of both the case group and the control group were greater than 0.05 in umbilical cord torsion, while in weeks 28-40, the weekly umbilical artery Doppler parameters were less than 0.05, with statistical significance, as shown in Table 2.

**Table 2.** Logistic regression analysis of S/D, RI, PI and PSV of umbilical artery between the case group and the control group at 21-40 weeks of gestation.

| GA | S/D |  | RI |  | PI |  | PSV |  |
| --- | --- | --- | --- | --- | --- | --- | --- | --- |
|  | OR(95%CL) | P-value | OR(95%CL) | P-value | OR(95%CL) | P-value | OR(95%CL) | P-value |
| 21 | 0.324(0.139-0.753) | 0.009 | 0.000(0.000-0.030) | 0.005 | 0.016(0.001-0.246) | 0.003 | 0.944(0.877-1.015) | 0.120 |
| 22 | 0.815(0.520-1.277) | 0.372 | 2.725(0.054-137.377) | 0.616 | 0.886(0.183-4.292) | 0.881 | 1.010(0.970-1.051) | 0.631 |
| 23 | 0.768(0.461-1.277) | 0.308 | 0.191(0.002-20.673) | 0.489 | 0.287(0.043-1.920) | 0.198 | 0.981(0.941-1.023) | 0.378 |
| 24 | 0.721(0.338-1.539) | 0.397 | 0.045(0.000-39.299) | 0.370 | 0.212(0.011-4.122) | 0.306 | 0.985(0.912-1.065) | 0.705 |
| 25 | 0.234(0.044-1.256) | 0.900 | 0.000(0.000-27.335) | 0.158 | 0.008(0.000-1.839) | 0.820 | 0.913(0.803-1.037) | 0.161 |
| 26 | 0.210(0.043-1.024) | 0.054 | 0.000(0.000-1.847) | 0.065 | 0.012(0.000-1.224) | 0.061 | 1.048(0.938-1.170) | 0.405 |
| 27 | 0.194(0.037-1.004) | 0.051 | 0.000(0.000-1.335) | 0.057 | 0.012(0.000-1.344) | 0.066 | 0.909(0.819-1.009) | 0.074 |
| 28 | 0.248(0.082-0.746) | 0.013 | 0.001(0.000-0.587) | 0.034 | 0.036(0.002-0.589) | 0.020 | 0.927(0.868-0.989) | 0.021 |
| 29 | 0.276(0.109-0.701) | 0.007 | 0.002(0.000-0.580) | 0.031 | 0.065(0.005-0.920) | 0.043 | 0.952(0.910-0.996) | 0.032 |
| 30 | 0.210(0.101-0.436) | 0.000 | 0.000(0.000-0.016) | 0.000 | 0.046(0.008-0.277) | 0.001 | 0.964(0.932-0.997) | 0.034 |
| 31 | 0.355(0.185-0.680) | 0.002 | 0.005(0.000-0.206) | 0.005 | 0.141(0.024-0.842) | 0.032 | 0.942(0.911-0.975) | 0.001 |
| 32 | 0.354(0.178-0.705) | 0.003 | 0.005(0.000-0.264) | 0.009 | 0.112(0.019-0.673) | 0.017 | 0.928(0.895-0.962) | 0.000 |
| 33 | 0.345(0.129-0.927) | 0.035 | 0.002(0.000-0.430) | 0.024 | 0.057(0.005-0.690) | 0.024 | 0.887(0.838-0.938) | 0.000 |
| 34 | 0.085(0.023-0.311) | 0.000 | 0.000(0.000-0.025) | 0.001 | 0.009(0.001-0.145) | 0.001 | 0.945(0.903-0.989) | 0.014 |
| 35 | 0.262(0.098-0.699) | 0.007 | 0.006(0.000-0.687) | 0.034 | 0.056(0.005-0.598) | 0.017 | 0.955(0.915-0.997) | 0.036 |
| 36 | 0.289(0.152-0.550) | 0.000 | 0.011(0.001-0.194) | 0.002 | 0.087(0.020-0.372) | 0.001 | 0.954(0.928-0.980) | 0.001 |
| 37 | 0.424(0.262-0.687) | 0.000 | 0.053(0.006-0.495) | 0.010 | 0.230(0.080-0.661) | 0.006 | 0.958(0.940-0.976) | 0.000 |
| 38 | 0.378(0.223-0.642) | 0.000 | 0.043(0.004-0.449) | 0.009 | 0.180(0.058-0.557) | 0.003 | 0.949(0.928-0.971) | 0.000 |
| 39 | 0.408(0.244-0.682) | 0.001 | 0.038(0.004-0.354) | 0.004 | 0.231(0.079-0.678) | 0.008 | 0.936(0.915-0.958) | 0.000 |
| 40 | 0.407(0.177-0.935) | 0.034 | 0.028(0.001-0.876) | 0.042 | 0.097(0.014-0.673) | 0.018 | 0.934(0.991-0.999) | 0.011 |

OR, odds ratio; CL, confidence interval; GA, Gestational age; PSV: peak systolic velocity; S/D: systolic/diastolic ratio; RI: resistance index; PI: pulsatility index

## 4. Discussion

This was a single-center retrospective case-control study. The results showed that the peak velocity of umbilical artery contraction (PSV) was positively correlated with gestational age in both the case group and the control group, and the values of the umbilical artery Doppler resistance related parameters (S/D, RI, PI) were negatively correlated with gestational age.

The results of this study showed that there was no significant difference in the mean value of umbilical artery Doppler parameters between the case group and the control group at 21-27 weeks of pregnancy (P > 0.05). The values of umbilical artery Doppler parameters in the 28-40 weeks gestation group were lower than those in the control group, and Logistic regression analysis showed that the difference was significant (P < 0.05). We hypothesize that umbilical cord torsion may begin in early or second trimester, and then gradually increase with gestational age. During the second trimester, the protective structure of the umbilical cord gives it some resistance to excessive torsion^[1-2]^, the changes of umbilical artery blood flow mechanics were not obvious, and the difference of Doppler parameters was not statistically significant. However, with the gradual aggravation of torsion in late pregnancy, the changes of umbilical artery blood flow mechanics were obvious, and the difference of Doppler parameters was statistically significant.

When the arteries in other parts of the human body are narrowed, the Doppler changes of the artery blood flow at the distal end of the stenosis are low-speed and low-pulsation changes, forming a “small slow wave”^[12,13]^. We believe that the Doppler changes of umbilical artery blood flow during umbilical cord torsion are consistent with the changes of blood flow Doppler changes after arterial stenosis in other parts:because umbilical cord torsion occurs mostly at the umbilical wheel^[3]^, the umbilical artery at the umbilical wheel is narrowed by the torsion of the umbilical cord,resulting in “small slow wave” changes in the blood flow Doppler of the umbilical artery at the distal end of the umbilical wheel (Fig. 1).

**Fig. 1.**
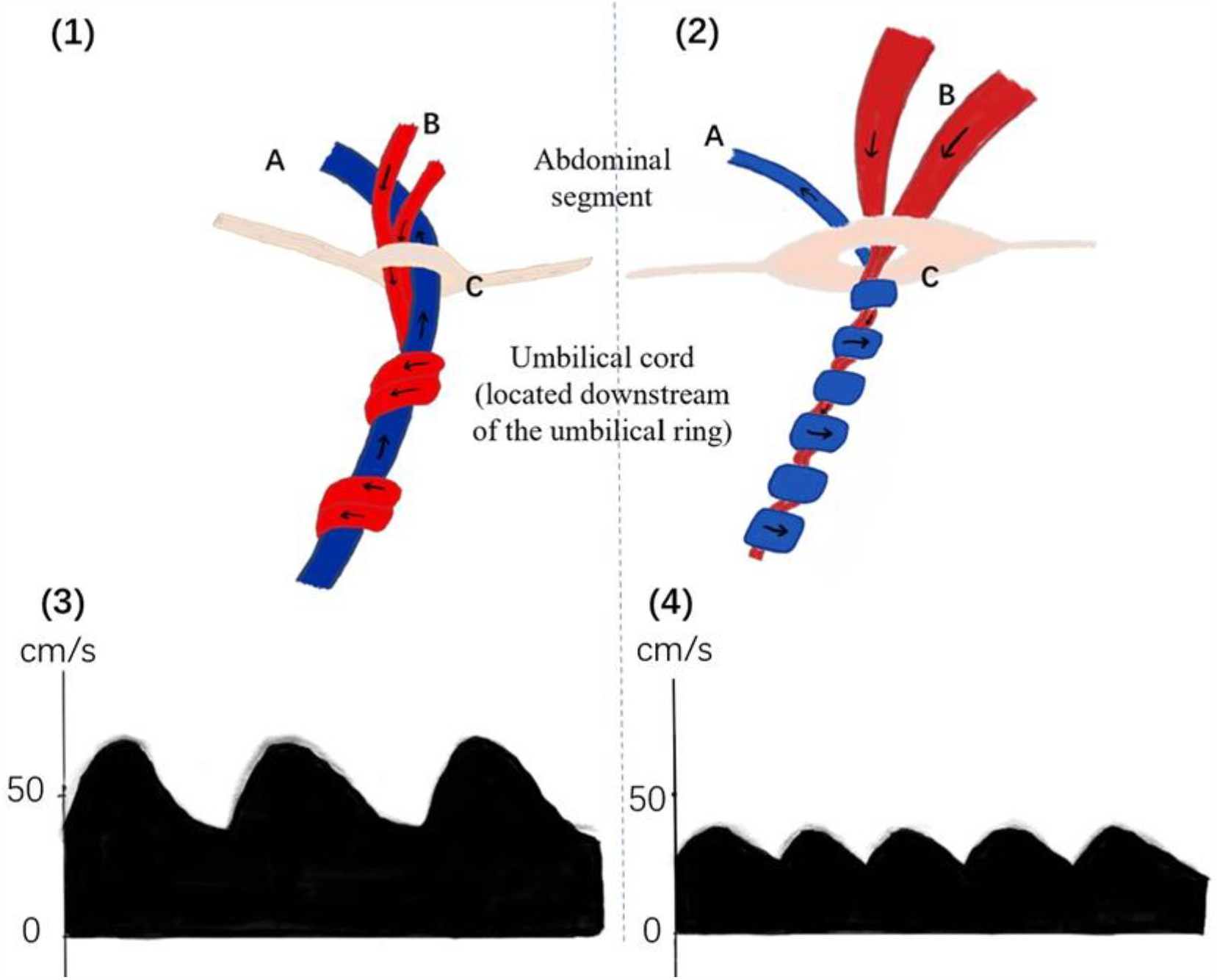
The umbilical cord (located downstream of the umbilical ring). (1) When the normal umbilical cord passes through the umbilical ring, there is no stenosis of the umbilical artery; (2) When the umbilical cord is twisted, the umbilical artery at the umbilical ring is narrowed due to torsion, and the peak systolic velocity and resistance of the umbilical artery downstream of the umbilical ring are reduced. (3) Normal umbilical artery Doppler waveform; (4) When the umbilical cord is twisted, the umbilical artery changes with low velocity and low resistance. A: umbilical vein; B: umbilical artery; C: the umbilical ring.

The Doppler parameters of umbilical artery blood flow are the routine indexes of prenatal ultrasound examination. At present, most of the bad pregnancies are related to the abnormal increase of umbilical artery blood Doppler resistance parameters^[14-18]^. When obstetricians suspect fetal distress or abnormal fetal movement, they usually pay more attention to the abnormal increase in Doppler resistance of umbilical artery blood flow. If there is no abnormal increase in Doppler resistance of umbilical artery blood flow, we often relax our vigilance and think that the fetus is safe for the time being, and it is recommended that the fetus be reexamined in the near future. During the time waiting for reexamination, the fetus may die in the uterus due to torsion of the umbilical cord, missing the best rescue opportunity. Therefore, when the fetus has abnormal fetal heart monitoring and abnormal fetal movement, and the Doppler parameters of umbilical artery blood flow do not increase abnormally or decrease instead, the possibility of umbilical cord torsion should be considered. It has been reported that the high helix of umbilical cord is related to umbilical cord torsion^[19-21]^. It has been reported that when the umbilical cord is high helical, the umbilical vein blood flow velocity at the umbilical wheel will increase^[22,23]^ or the a-wave reverse of the venous catheter will occur^[24]^. Therefore, when suspected umbilical cord torsion, we also need to pay attention to the contents of unconventional ultrasound examination, such as umbilical cord helix, umbilical vein flow velocity and venous catheter spectrum and so on.

Next, we share a case. At 32 weeks of pregnancy, the pregnant woman realized that the fetal movement was normal, and the fetal heart monitoring in the other hospital showed a poor response. She was referred to our hospital for further examination. The previous ultrasound examination showed that the double umbilical artery was found to be a single umbilical artery.

The PSV of umbilical artery was 36.42 cm/s,S/D and 1.84 Magi was 0.46 and Pi was 0.60, in which PSV was lower than normal 5%th^[25]^, and Spicer D, RI and PI were lower than 10%th^[25]^(Fig. 2A). Fetal heart rate monitoring indicates that the response is poor and the related parameters of Doppler resistance of umbilical artery blood flow are low, suggesting that there may be umbilical cord torsion. Therefore, we examined the shape of the umbilical cord and the blood flow of the venous catheter: it was found that the fetal umbilical cord spirally increased, UCI0.76^[26]^ (Fig. 2B,2C), and the umbilical vein blood flow velocity at the umbilical cord wheel (PSV 69cm/s) was higher than that of 95%th^[23]^(Fig.2D). The a-wave notch of fetal venous duct was significantly deepened and PI increased, which was larger than that of 95%th^[25]^(Fig.2E). In summary, we considered the possibility of umbilical cord torsion, and the obstetrician performed a cesarean section to terminate the pregnancy. The postoperative diagnosis confirmed umbilical cord torsion at 36 weeks (Fig.2F). Postoperative pathology of the placental adnexa revealed two umbilical arteries (one of which was atresia) and one umbilical vein.

**Fig. 2.**
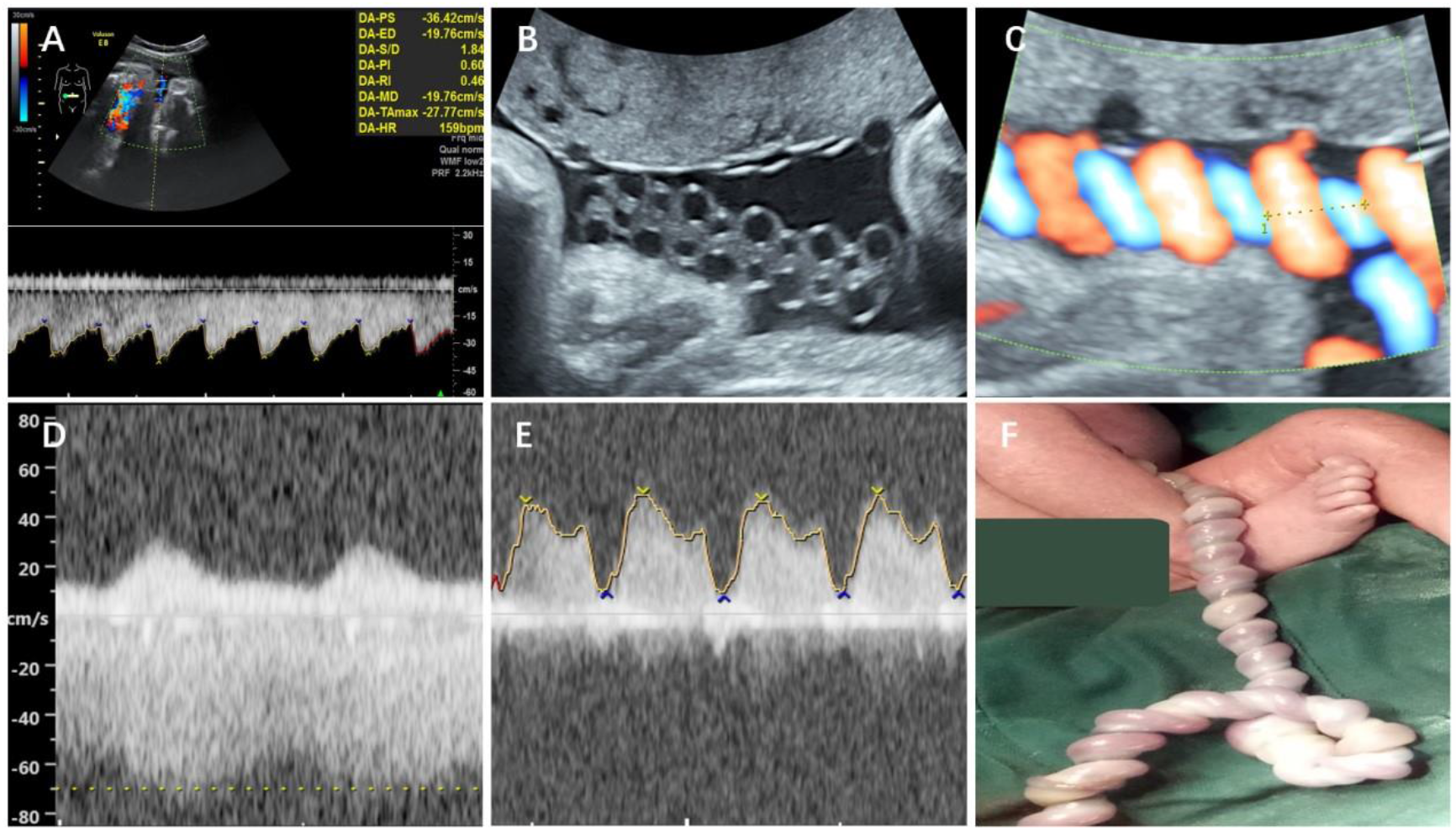
A case of umbilical cord torsion. A: Abnormal fetal heart monitoring at 32 weeks of gestation, frequency spectrum of umbilical cord free segment. PSV is lower than 5th%, S/D, RI, PI is lower than 10th%. B: 2D image of high spiral umbilical (changing like chain); C: Color Doppler image of high spiral umbilical (UCI 0.76); D: The umbilical vein flow velocity at the umbilical ring increased (PSV 69cm/s); E: Doppler spectrum of intravenous catheter (a notch significantly deepened, PI 1.22). F: Umbilical cord torsion > 30 weeks after birth.

## 5. Conclusions

The decrease of umbilical artery Doppler parameters in late pregnancy is significantly correlated with umbilical cord torsion, which may be used as a clue for prenatal ultrasound screening of umbilical cord torsion. The Doppler characteristics of umbilical artery blood flow during umbilical cord torsion are consistent with the basic principle of the changes of blood flow Doppler parameters after vascular stenosis in other parts of the human body. A multicenter prospective cohort study is needed in the future.

## Data Availability

All data produced in the present study are available upon reasonable request to the authors

## Acknowledgment

Not applicable.

## Abbreviations

GA: gestational age
PSV: peak systolic velocity
S/D: systolic/diastolic ratio
RI: resistance index
PI: pulsatility index

